# Optimizing Stem Cell Infusion Timing in the Prevention of Acute Graft versus Host Disease

**DOI:** 10.1101/2024.02.06.24302168

**Authors:** Yiwen Hou, Yue Wu, Zhonglin Zhang, Liang Wang, Zhiwei Liu, Baolin Tang, Kaidi Song, Guangyu Sun, Xiaoyu Zhu, Cheng Zhan

## Abstract

Allogeneic hematopoietic stem cell transplantation (allo-HSCT) is a cornerstone treatment for a broad spectrum of malignant and nonmalignant hematological disorders. However, the success of allo-HSCT is often overshadowed by acute graft-versus-host disease (aGVHD), a life-threatening complication with limited preventive options. Here, we found that the incidence and severity of aGVHD after allo-HSCT are highly dependent on the circadian timing of stem cell infusion. The incidence rate of aGVHD in patients decreased by approximately 50% for early infusion (before 2:00 pm) compared to later infusion (after 2:00 pm). Early-infused patients also experienced significantly lower three-year transplant-related mortality and improved GVHD-free, relapse-free survival. Animal studies using an aGVHD mouse model show that this improvement is mainly due to the recipient’s rhythm rather than the donor’s. Mechanistically, compared with late infusions, early infusions significantly reduced the levels of the pro-inflammatory cytokine IL-1α following the conditioning regimen and subsequently suppressed T-cell activation and differentiation after transplantation. Our study suggests that scheduling stem cell infusions early in the day could be a simple yet transformative intervention for the prevention of aGVHD.

## INTRODUCTION

A primary challenge in allo-HSCT is graft-versus-host disease (GVHD), stemming from a potent immunologic assault on the recipient’s organs or tissues, which accounts for a considerable proportion of transplant-related mortality post-transplant(Blazar et al., 2012; Ferrara et al., 2009). Current preventative strategies for GVHD include the use of calcineurin inhibitors (CNIs), antimetabolites, and in vivo T cell depletion with anti-thymocyte globulin (ATG)(Bacigalupo et al., 2001; Bolanos-Meade et al., 2023; Ratanatharathorn et al., 1998; Storb et al., 1986; Zeiser and Blazar, 2017). Despite these effective prophylactic measures, 30% to 50% of patients still suffer from acute GVHD (aGVHD) after allogeneic transplantation(Zeiser and Blazar, 2017). Moreover, these interventions can also dampen the beneficial graft-versus-leukemia effect, increasing the risk of infection and disease relapse(Blazar et al., 2012; Shlomchik, 2007). Consequently, there is an ongoing imperative to reduce both the incidence and severity of aGVHD after allo-HSCT.

The circadian rhythm has emerged as a crucial regulator of immune functions and immune-mediated diseases(Scheiermann et al., 2013). The circadian rhythm can significantly influence various aspects of the hematopoietic system, including the yield, release, migration, and engraftment potential of stem cells(Aardal and Laerum, 1983; Abrahamsen et al., 1998; D’Hondt et al., 2004; Lucas et al., 2008; Mendez-Ferrer et al., 2008). However, the implications of circadian rhythms for aGVHD management and patient outcomes in allo-HSCT patients have not been explored. The present protocols for allogeneic transplantation do not typically account for circadian considerations. In this study, we conducted hybrid research involving both allo-HSCT patients and animal models to investigate the impact of the timig of day of stem cell infusion on aGVHD. Our findings underscore that the timing of stem cell infusion profoundly influences the incidence and severity of aGVHD following allo-HSCT.

## RESULTS

### The diagram of the clinical research and patient characteristics

From January 2015 to May 2023, 230 consecutive patients who underwent unmanipulated peripheral blood stem cell transplantation (PBSCT) at Anhui Provincial Hospital were screened. All patients received mobilized peripheral blood stem cells as a graft source. For patients who had no matched identical sibling donors or matched unrelated donors, haplo-donors were considered. Patients who received their second transplantation were excluded (n = 26). A total of 204 patients who underwent PBSCT using an identical sibling donor (ISD) or haplo-identical donor (HID) were ultimately enrolled in the analysis **(Figure S1)**.

The patients were categorized based on the stem cell infusion time, which was recorded as the beginning of infusion. The timing of stem cell infusion ranged between 11:00 am and 10:00 pm, and the median time for the whole cohort was 2:00 pm. Patients who received infusions earlier than 2:00 pm were considered the early-infused group (n = 97) with a 1:00 pm median time, and those who received infusions later than or equal to 2:00 pm were considered the late-infused group (n = 107) with a 5:00 pm median time. **Table S1** lists the patient demographics and transplantation features, and the baseline characteristics were comparable between the early-infused group (before 2:00 pm) and the late-infused group (after 2:00 pm).

### Effect of the infusion timing on GVHD

The cumulative incidence of grade II-IV and III-IV aGVHD by day 100 after allo-HSCT was significantly lower (20.6% [95% *CI*, 13.2-29.2%] vs. 38.3% [95% *CI*, 29.1-47.5%], *P* = 0.009 for grade II-IV aGVHD; 9.3% [95% *CI*, 4.5-16.1%] vs. 27.1% [95% *CI*, 19.0-35.8%], *P* < 0.001 for grade III-IV aGVHD) in the early-infused group (**Figure 1)**. In the early-infused group, 20 patients experienced Grade II-IV aGVHD; among these, 11 met the criteria for Grade II aGVHD (8 of them with gastrointestinal involvement and 4 with cutaneous involvement), the other 9 patients developed Grade III-IV aGVHD (9 with gastrointestinal involvement, 4 with hepatic involvement and 1 with cutaneous involvement), and 2 patients died due to uncontrolled aGVHD. In contrast, the late-infused group had 41 patients with Grade II-IV aGVHD, and 12 of those met the criteria for Grade II aGVHD (4 of them with gastrointestinal involvement and 8 with cutaneous involvement). A significant portion, 29 patients, progressed to grade III-IV aGVHD (28 with gastrointestinal involvement, 7 with hepatic involvement and 6 with cutaneous involvement), and there were 7 deaths attributable to uncontrolled aGVHD. None of the patients in either circadian infusion group met the criteria for late-onset aGVHD. These results clearly demonstrated a markedly lower incidence and severity of aGVHD in the early-infused group than in the late-infused group.

**Fig. 1.**
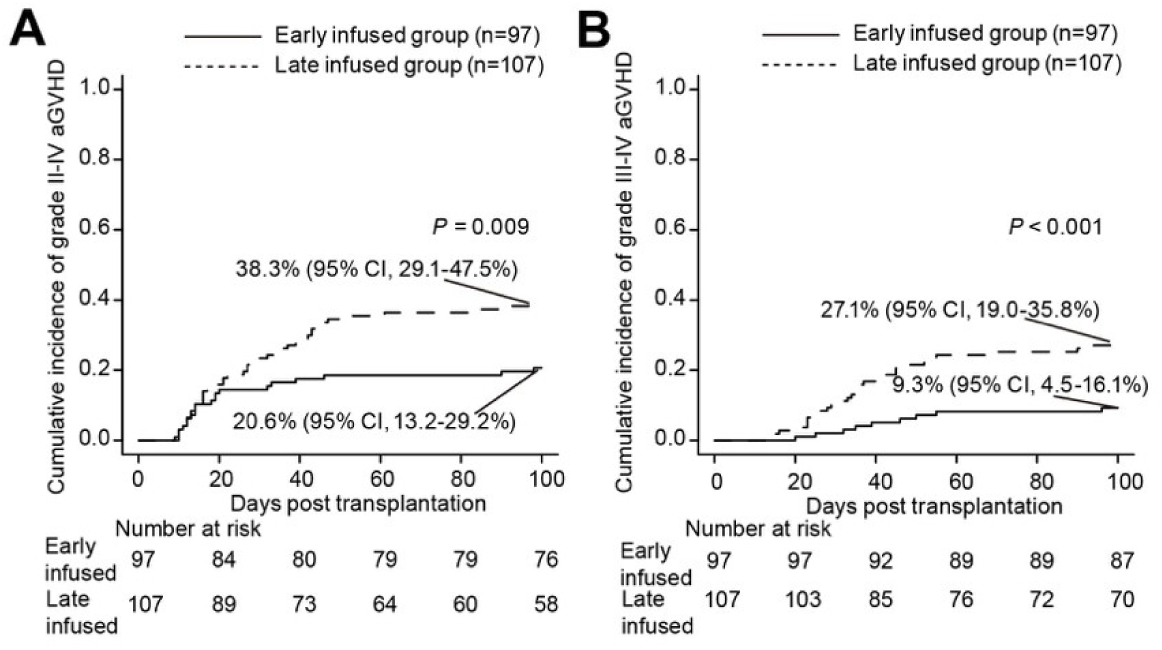
Cumulative incidences of grade II-IV aGVHD (A) and grade III-IV aGVHD (B) in the early-infused group and late-infused group.

To further analyze whether the circadian timing of stem cell infusion is a risk factor for aGVHD, we performed both univariate and multivariate regression analyses considering variables such as age, recipient sex, disease diagnosis (malignant vs. benign), donor/recipient sex compatibility, HLA antibody, ABO compatibility, donor type (ISD vs. HID), conditioning regimen, GVHD prophylaxis regimen, counts of infused total nucleated cells (TNC), counts of infused CD34^+^ cells, usage of MTX, and occurrence of engraftment syndrome (ES) and timing of stem cell infusion. As shown in **Table 1**, the univariate analysis indicated that the development of Grade II-IV aGVHD was significantly associated with stem cell infusion timing (hazard ratio (HR) = 1.13, 95% *CI*, 1.04-1.23, *P* = 0.005). All variables with a *P* value < 0.2 were included in the multivariate regression analysis, which identified stem cell infusion timing (HR = 1.13, 95% *CI*, 1.04-1.23, *P* = 0.005) as the sole independent risk factor for Grade II-IV aGVHD. Moreover, stem cell infusion timing was also a distinct risk factor for the development of Grade III-IV aGVHD according to both univariate and multivariate analyses (HR = 1.16, 95% *CI*, 1.05-1.27, *P* = 0.003 in univariate analysis, HR = 1.16, 95% *CI*, 1.05-1.27, *P* = 0.003 in multivariate analysis). Consistent with the findings of previous reports(Flowers et al., 2011; Jagasia et al., 2012), another independent risk factor for Grade III-IV aGVHD was D/R sex compatibility (female to male) (HR = 2.34, 95% *CI*, 1.24-4.42, *P* = 0.01 in univariate analysis, HR = 2.32, 95% *CI*, 1.24-4.35, *P* = 0.009 in multivariate analysis). These findings underscore the significance of stem cell infusion timing as a pivotal risk factor for aGVHD in PBSCT patients.

**Table 1.** Univariate and multivariate analyses of the factors associated with acute GVHD.

| Variable | Univariate analysis |  |  | Multivariate analysis |  |  |
| --- | --- | --- | --- | --- | --- | --- |
| | HR | 95%CI | $P$ | HR | 95%CI | $P$ |
| Grade II-IV acute GVHD |  |  |  |  |  |  |
| Age, y | 1.01 | 0.99-1.03 | 0.210 |  |  |  |
| Recipient sex |  |  |  |  |  |  |
| Female | 0.68 | 0.40-1.14 | 0.140 | 0.61 | 0.36-1.05 | 0.075 |
| Diagnose |  |  |  |  |  |  |
| Malignant | 1.45 | 0.70-3.03 | 0.320 |  |  |  |
| D/R sex |  |  |  |  |  |  |
| Female to Male | 1.35 | 0.80-2.27 | 0.260 |  |  |  |
| HLA antibody |  |  |  |  |  |  |
| Positive | 0.99 | 0.51-1.91 | 0.980 |  |  |  |
| ABO compatibility | 1.13 | 0.87-1.45 | 0.370 |  |  |  |
| Donor type |  |  |  |  |  |  |
| ISD | 1.21 | 0.70-2.08 | 0.50 |  |  |  |
| Conditioning regimen |  |  |  |  |  |  |
| RIC | 0.88 | 0.48-1.64 | 0.69 |  |  |  |
| GVHD Prophylaxis regimen |  |  |  |  |  |  |
| without rATG or PT-Cy | 1 |  |  |  |  |  |
| with rATG only | 0.89 | 0.37-2.16 | 0.790 |  |  |  |
| with PT-Cy only | 0.74 | 0.34-1.61 | 0.450 |  |  |  |
| with rATG and PT-Cy | 0.90 | 0.37-2.21 | 0.820 |  |  |  |
| Infused TNC count | 1.01 | 0.98-1.03 | 0.710 |  |  |  |
| Infused CD34<sup>+</sup> cell count | 0.97 | 0.94-1.01 | 0.180 | 0.97 | 0.93-1.01 | 0.170 |
| MTX prophylaxis | 0.74 | 0.31-1.73 | 0.480 |  |  |  |
| Engraftment syndrome | 1.78 | 0.86-3.69 | 0.120 | 1.85 | 0.87-3.93 | 0.110 |
| Infusion time | 1.13 | 1.04-1.23 | 0.005 | 1.13 | 1.04-1.23 | 0.005 |
| Grade III-IV acute GVHD |  |  |  |  |  |  |
| Age, y | 1.00 | 0.98-1.02 | 0.860 |  |  |  |
| Recipient sex |  |  |  |  |  |  |
| Female | 0.75 | 0.39-1.43 | 0.140 | 1.14 | 0.45-2.90 | 0.780 |
| Diagnose |  |  |  |  |  |  |
| Malignant | 1.37 | 0.53-3.55 | 0.520 |  |  |  |
| D/R sex |  |  |  |  |  |  |
| Female to Male | 2.34 | 1.24-4.42 | 0.01 | 2.32 | 1.24-4.35 | 0.009 |
| HLA antibody |  |  |  |  |  |  |
| Positive | 0.81 | 0.32-2.02 | 0.650 |  |  |  |
| ABO compatibility | 1.09 | 0.80-1.50 | 0.580 |  |  |  |
| Donor type |  |  |  |  |  |  |
| ISD | 1.21 | 0.60-2.43 | 0.60 |  |  |  |
| Conditioning regimen |  |  |  |  |  |  |
| RIC | 0.89 | 0.40-1.96 | 0.77 |  |  |  |
| GVHD Prophylaxis regimen |  |  |  |  |  |  |
| without rATG or PT-Cy | 1 |  |  |  |  |  |
| with rATG only | 0.69 | 0.20-2.30 | 0.540 |  |  |  |
| with PT-Cy only | 0.70 | 0.25-1.97 | 0.500 |  |  |  |
| with rATG and PT-Cy | 1.26 | 0.43-3.67 | 0.670 |  |  |  |
| Infused TNC count | 1.01 | 0.99-1.04 | 0.250 |  |  |  |
| Infused CD34<sup>+</sup> cell count | 0.96 | 0.89-1.02 | 0.170 | 0.97 | 0.91-1.03 | 0.260 |
| MTX prophylaxis | 1.05 | 0.39-2.85 | 0.920 |  |  |  |
| Engraftment syndrome | 1.85 | 0.75-4.54 | 0.180 | 2.06 | 0.80-5.31 | 0.140 |
| Infusion time | 1.16 | 1.05-1.27 | 0.003 | 1.16 | 1.05-1.27 | 0.003 |

GVHD, graft-versus-host disease; D/R, donor/recipient; HLA, human leucocyte antigen; ISD, identical sibling donor; rATG, rabbit anti-thymocyte globulin; PT-Cy, post-transplant cyclophosphamide; TNCs, total nucleated cells; MTX, methotrexate

The cumulative incidences of 3-year overall and moderate-to-severe chronic GVHD (cGVHD) did not significantly differ between the two groups (*P* = 0.476 and 0.108, respectively) (**Figure S2**). In the early-infused group, 13 patients developed moderate to severe cGVHD, while 24 patients were affected in the late-infused group. The predominant manifestation among these was sclerodermatous skin changes, present either as an isolated symptom or concomitant with pulmonary, hepatic, or gastrointestinal tract involvement.

### Effect of circadian rhythm on other transplantation outcomes

While the probabilities of 3-year overall survival (OS) and disease-free survival (DFS) were not significantly different between the two groups (early-infused group vs. late-infused group: OS, 74.7% [95% *CI*, 64.4-82.5%] vs. 65.5% [95% *CI*, 55.5-73.8%]; DFS, 70.9% [95% *CI*, 60.4-79.1%] vs. 61.6% [95% *CI*, 51.5-70.2%]; *P* = 0.068, 0.097, respectively), the probability of 3-year GVHD free, relapse-free survival (GRFS) was significantly greater in the early-infused group than in the late-infused group (52.9% [95% *CI*, 42.4-62.4%] vs. 33.9% [95% *CI*, 25.0-43.0%], *P* < 0.001) (**Figure 2**). These results suggest that stem cell infusion early in the day could significantly improve long-term GRFS after allo-HSCT.

**Fig. 2.**
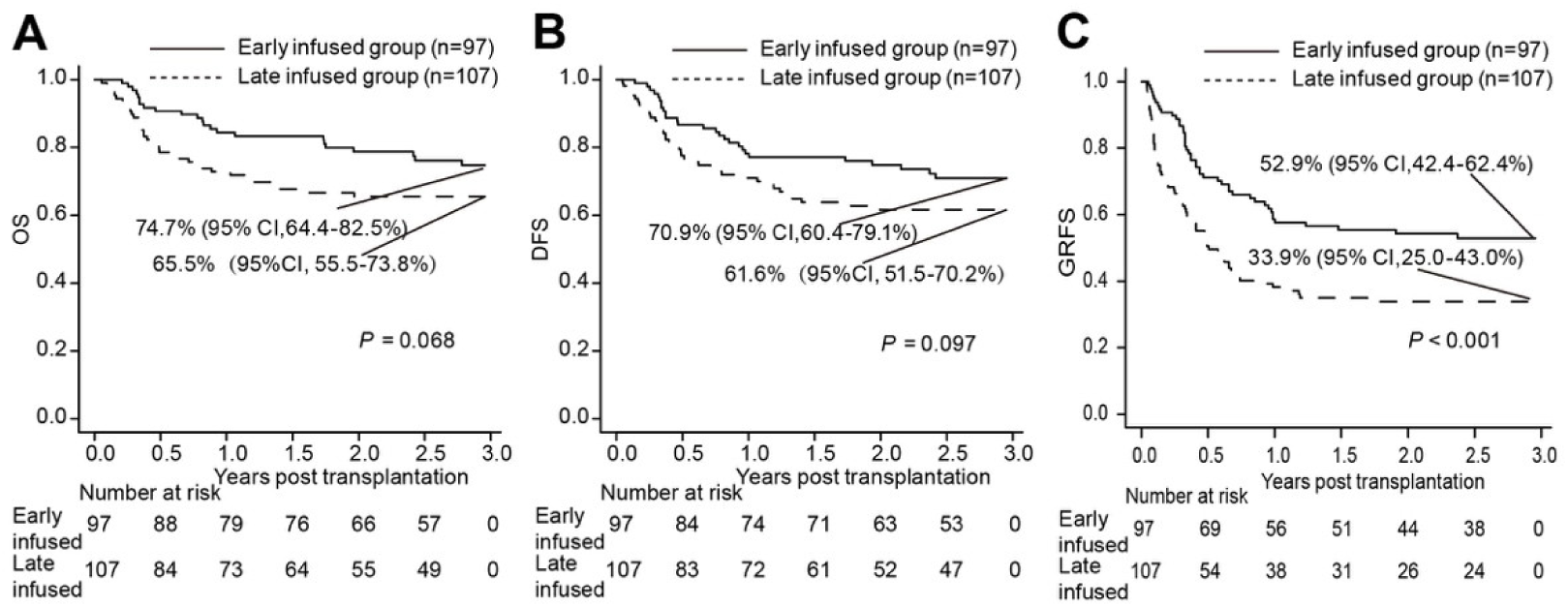
Transplantation outcomes between the early-infused patients and late-infused patients. **(A)** Probability of overall survival (OS). **(B)** Probability of disease-free survival (DFS). **(C)** Probability of graft-versus-host disease (GVHD) free, relapse-free survival (GRFS).

Among all the patients, 1 had primary graft failure (PGF), and 1 died in the early phase posttransplantation. The cumulative incidence of neutrophil engraftment by day 30 was lower in the late-infused group compared to the early-infused group (98.1% [95% *CI*, 91.3-99.6%] vs. 100.0%, P =0.048) (**Figure S3A**). One patient in the early-infused group and 5 patients in the late-infused group died before day 100 as a result of transplant-related mortality (TRM) without obtaining platelet recovery. The cumulative incidence of platelet recovery by day 100 was lower in the late-infused group compared to the early-infused group (83.3% [95% *CI*, 74.5-89.3%] vs. 92.8% [95% *CI*, 85.1-96.6%], *P* =0.029) (**Figure S3B**). In general, early infusions promote the hematopoietic recovery of patients post transplantation.

We then compared complications post transplantation between the two groups. Other transplant-related outcomes, such as the incidence of hepatic veno-occlusive disease (HVOD), cytomegalovirus (CMV) antigenemia, hemorrhagic cystitis (HC), bacterial septicemia, and engraftment syndrome (ES), did not significantly differ between the two groups (*P* > 0.05). As shown in **Table S2**, the stem cell infusion timing did not have a marked impact on other complications after transplantation.

The cumulative incidence of 3-year TRM was significantly lower in the early-infused group (13.8% [95% *CI*, 7.7-21.7%] vs. 24.8% [95% *CI*, 16.9-33.4%], *P* = 0.041), while the incidence of relapse did not significantly differ between the two groups (late-infused group vs. early-infused group, 13.6% [95% *CI*, 7.8-21.0%] vs. 15.2% [95% *CI*, 8.7-23.4%] *P* = 0.843) (**Figure 3**). Twenty-three patients in the early-infused group and 37 patients in the late-infused group died (**Table S3)**. The primary causes of mortality in the early-infused group were relapse (52.2%), multiorgan dysfunction syndrome (MODS) (17.4%) and infection (13.0%). In the late-infused group, the primary causes were relapse (29.7%), infection (21.6%) and uncontrolled aGVHD (18.9%). Taken together, these results suggest that early infusions of stem cells significantly reduce TRM posttransplantation.

**Figure 3.**
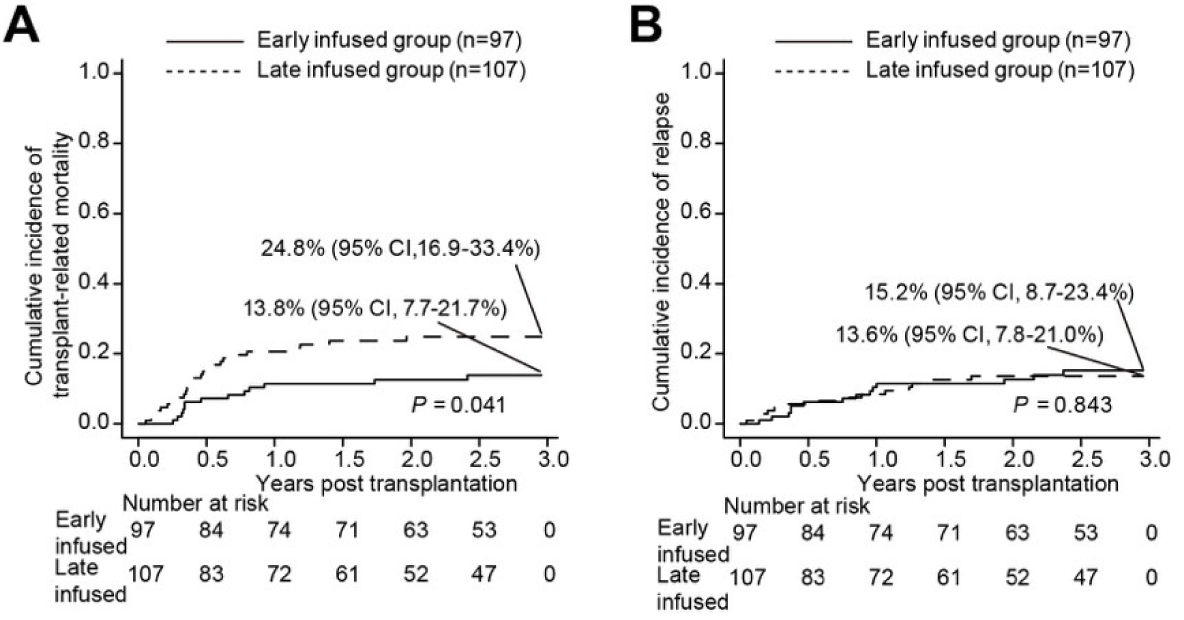
Cumulative incidences of 3-year transplant-related mortality (A) and relapse (B) in the early-infused group and late-infused group.

### Stem cell infusion timing impacts aGVHD in mice

To investigate the underlying mechanisms by which stem cell infusion timing affects aGVHD development and improves prognosis, we utilized an MHC-mismatched allo-HSCT mouse model in which bone marrow cells and splenocytes were transplanted from BALB/c (H2k^d^) mice into lethally irradiated C57BL/6J (H2k^b^) recipients (i.e., BALB/c→C57BL/6J) at either Zeitgeber Time 5 (i.e., ZT5) or ZT14 **(Figure 4A)**. As nocturnal animals, mice are inactive during ZT0 (light onset) to ZT12 and active from ZT12 (dark onset) to ZT24. Thus, ZT14 corresponds to early in the day in humans, while ZT5 corresponds late in the day.

**Fig. 4.**
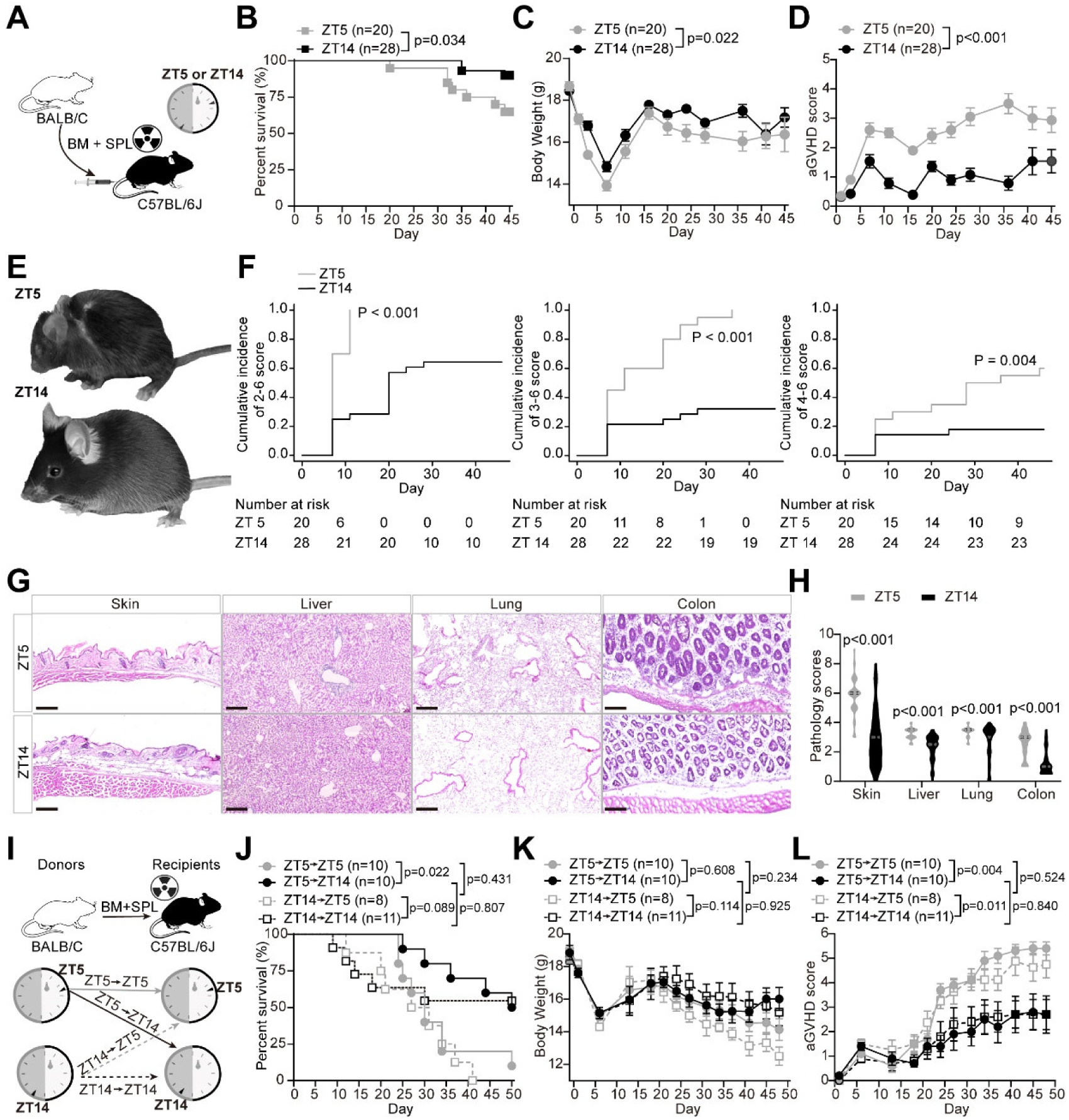
The impacts of stem cell infusion timing on aGVHD in mice. **(A)** Schematic of allo-HSCT in mice. C57BL/6J recipient mice at ZT5 and ZT14 received bone marrow cells and splenocytes (SPL) transplantation from BALB/c donor mice by tail vein injection following lethal total-body irradiation. **(B-D)** Survival rate (**B**), body weight (**C**), and aGVHD score (**D**) of lethally irradiated recipients after allo-HSCT. Data shown derive from three independent experiments. **(E)** Representative examples showing phenotypic differences on day 45 after allo-HSCT from the ZT5 (top) and ZT14 (bottom) groups. **(F)** Cumulative incidences of aGVHD scores at 2-6 (left), 3-6 (middle) and 4-6 (right) in the ZT5 and ZT14 groups. **(G** and **H)** Histopathologic analysis of skin, liver, lung, and colon section on day 45 after allo-HSCT. Representative images of hematoxylin and eosin staining (**G**) and quantification results (n=5-7 independent mice per group combined from three replicate experiments) (**H**). Scale bars, 200 μm. **(I-L)** The effects of recipient’s and donor’s rhythms on aGVHD. **I**, Experimental schematic. Both ZT5 and ZT14 recipient mice received transplantation from ZT5 or ZT14 donor mice. Survival rate (**J**), body weight (**K**), and aGVHD score (**L**). Data are presented as means ± SEMs and analyzed by two-way ANOVA followed by Bonferroni post hoc test (**C**, **D**, **K** and **L**), unpaired *t* test (**H**) and log-rank test (**B** and **J**).

Interestingly, mice transplanted at ZT14 (ZT14 group) exhibited a significantly lower mortality rate **(Figure 4B)** and less severe weight loss **(Figure 4C)** than those transplanted at ZT5 (ZT5 group). Crucially, the ZT14 group also had significantly lower aGVHD scores **(Figure 4D, E)** and significantly lower cumulative incidences of aGVHD of 2-6, 3-6, and 4-6 scores **(Figure 4F)**. Histopathologic analyses further corroborated these findings, revealing decreased tissue injury and inflammation in the skin, lungs, colon, and liver in the ZT14 group compared to the ZT5 group **(Figure 4G, H)**. No significant differences were observed between the groups regarding donor chimerism rates **(Figure S4)**. Collectively, these findings indicate a lower incidence and severity of aGVHD in the ZT14 group relative to the ZT5 group.

### The Recipient’s Circadian Rhythm is Crucial

To ascertain whether aGVHD incidence is affected more by the circadian rhythmicity of the recipient or donor, we administered stem cells from donors at ZT5 or ZT14 to ZT5 and ZT14 recipients post-irradiation **(Figure 4I)**. Significant differences were observed in survival rates and aGVHD scores based on the recipients’ circadian rhythm, with ZT14 recipients demonstrating substantially lower aGVHD scores than their ZT5 counterparts, regardless of the donor’s circadian rhythm **(Figure 4J-L)**. Even when donor rhythmicity was altered to ZT1 and ZT9, ZT14 recipients consistently exhibited less severe aGVHD symptoms compared to ZT5 recipients **(Figure S5).** Conversely, modifications to only the donor’s circadian rhythm did not significantly alter survival rates, bodyweight, or aGVHD scores **(Figure 4J-L)**. Therefore, these results suggest that the influence of the infusion timing on aGVHD is mainly due to the recipient’s rhythm rather than the donor’s.

### Cytokine levels and T-cell activation

The pathophysiology of aGVHD consists of three phases(Ferrara et al., 2009; Schroeder and DiPersio, 2011): (1) initial production of danger signals, such as pro-inflammatory cytokines, due to the conditioning regimen; (2) donor T cell activation and cytokine release; and (3) resultant host tissue damage. Given the significant effect of stem cell infusion timing on aGVHD outcomes, we hypothesized that these effects predominantly occur during the early stages of aGVHD development. We began by examining cytokine levels in murine serum following irradiation. Of the thirteen cytokines assessed, only IL-1α was significantly lower in the ZT14 recipients than in the ZT5 recipients **(Figure 5A)**, and no significant differences were observed for the other cytokines. IL-1α is a classic damage-associated molecular pattern (DAMP) molecule produced in response to cell death and tissue damage(Malik and Kanneganti, 2018). These results suggest less pronounced tissue damage from irradiation in the ZT14 group.

**Fig. 5.**
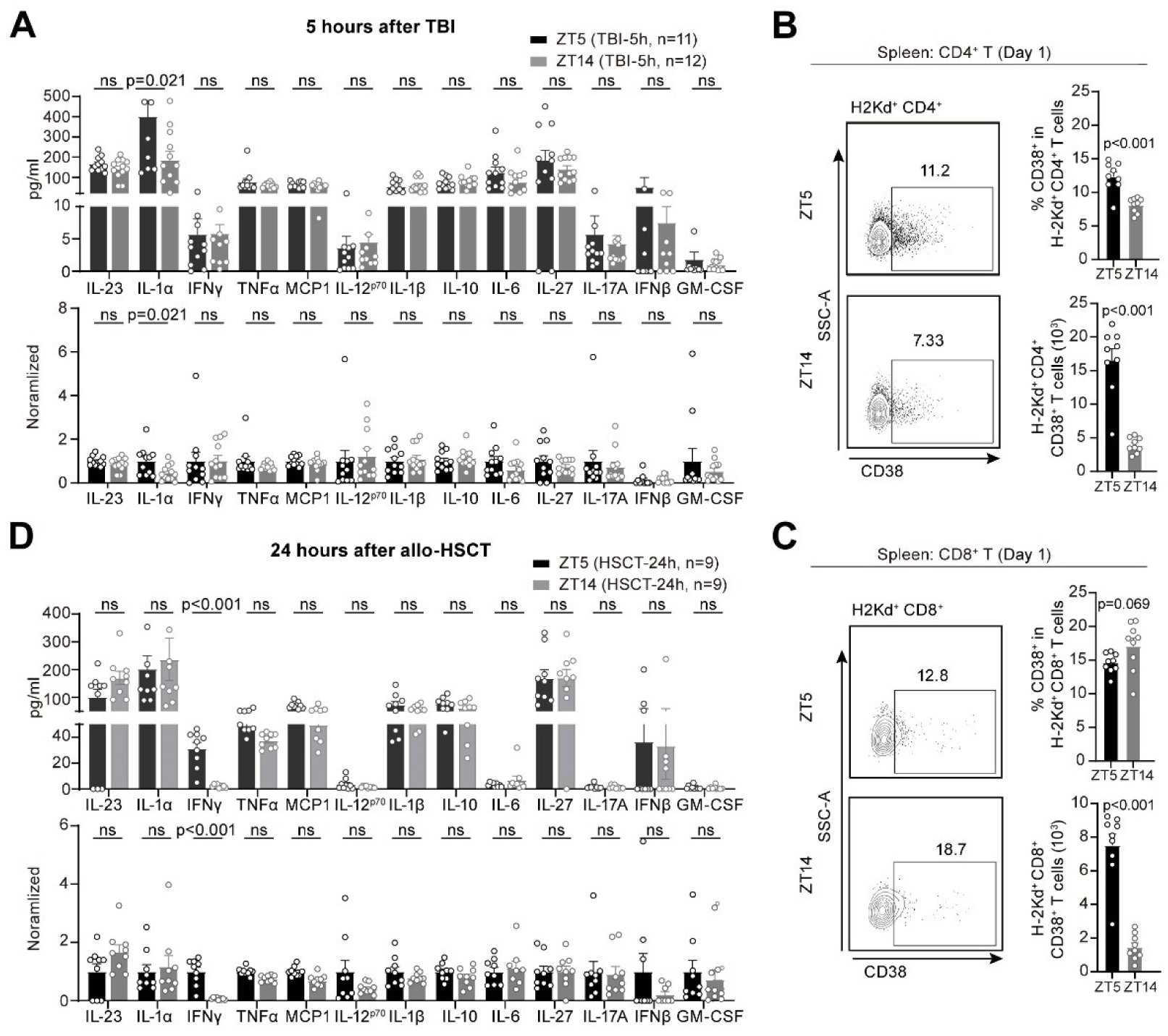
The impacts of the stem cell infusion timing on cytokine levels and T cell activation. **(A)** Absolute (top) and normalized (bottom) cytokine levels in the sera of recipient mice 5 hours after lethal irradiation. Mice were exposed to lethal irradiation at ZT0 and ZT9, with serum samples collected at ZT5 and ZT14 on the same day. For the bottom panel, cytokine levels in the ZT14 group were normalized to those in the ZT5 group for each cytokine. **(B** and **C)** Representative dot plots (left), proportions (right top) and absolute counts (right bottom) of donor activated (H2k^d+^ CD38^+^) CD4^+^ T cells (**B**) and CD8^+^ T cells (**C**) in the spleen of recipient mice 24 hours (day 1) after allo-HSCT. **(D)** Absolute (top) and normalized (bottom) cytokine levels in the sera of recipient mice 24 hours after allo-HSCT. Stem-cell infusions were performed allo-HSCT at ZT5 and ZT14, and serum samples were collected 24 hours later. For the bottom panel, cytokine levels in the ZT14 group were normalized to those in the ZT5 group for each cytokine. Data are presented as means ± SEMs and analyzed by unpaired *t* test. TBI, total body irradiation. ns, not significant.

IL-1α plays critical roles in inducing T cell activation(Ferrara et al., 1993). We then examined whether donor T cell activation was affected by the timing of stem cell infusion, using the activation marker CD38 **(Figure S6)**. At 24 hours (day 1) after allo-HSCT, both the percentages and absolute numbers of donor activated (H2k^d+^ CD38^+^) CD4^+^ T cells were significantly lower in the ZT14 group than in the ZT5 group **(Figure 5B)**. Although there was no significant difference in the percentage of donor activated (H2k^d+^ CD38^+^) CD8^+^ T cells between the two groups, the absolute number of donor activated CD8^+^ T cells was substantially lower in the ZT14 group than in the ZT5 group **(Figure 5C)**. IFNγ, released by activated T cells post-transplant, usually signals heightened T cell activation(Schroeder and DiPersio, 2011). In line with this, post-transplant IFNγ levels were significantly lower in the ZT14 group than in the ZT5 group **(Figure 5D)**. These findings indicate substantially decreased T cell activation in the ZT14 group compared with the ZT5 group after transplantation.

We further identified the differentiation state of the donor T cells by examining CD44 and CD62L expression. On day 1 after allo-HSCT, the percentage of naïve (CD44^-^ CD62L^+^) T cells was significantly greater in the ZT14 group than in the ZT5 group **(Figure 6A, C).** In contrast, the percentages of donor central memory (CD44^+^ CD62L^+^) and effector memory (CD44^+^ CD62L^-^) T cells (including both the CD4^+^ and CD8^+^ subsets) were significantly lower in the ZT14 group than in the ZT5 group **(Figure 6A, C)**. LikewiseSimilarly, the percentage of donor effector memory T cells was significantly lower in the ZT14 group than in the ZT5 group 7 days after allo-HSCT **(Figure 6B, D)**. Effector memory T cells can quickly produce cytokines and mediate immune responses against host tissue, which contributes to the pathology of aGVHD. Thus, these results suggest that early infusion (ZT14) may decrease the proportion of effector memory T cells, potentially creating an immune environment less conducive to the development of the harmful immune responses associated with aGVHD.

**Fig. 6.**
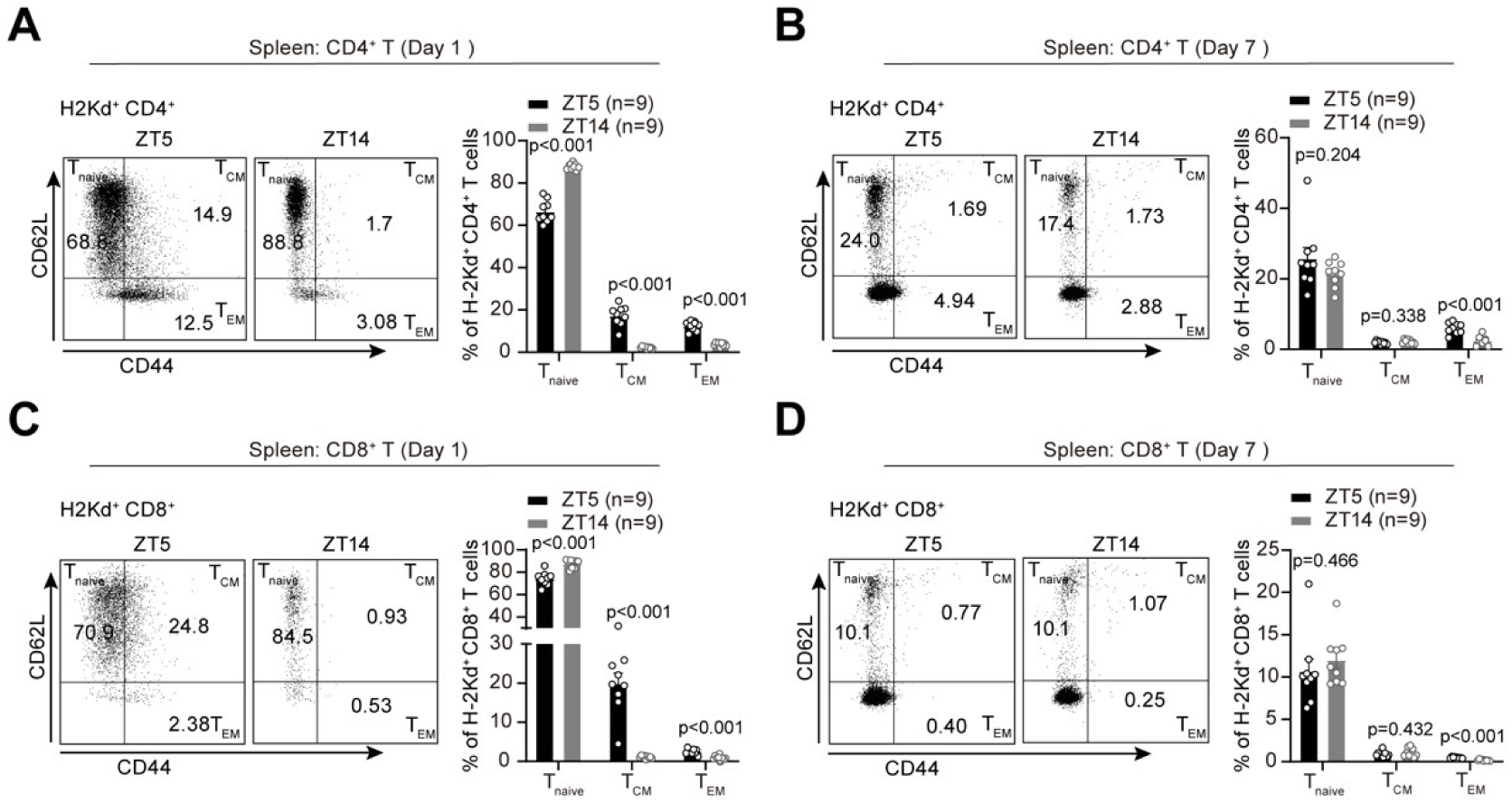
Impacts of stem cell infusion timing on the status of T cells. (**A** and **B**) Representative dot plots (left) and proportions (right) of CD4^+^ T_naive_, T_CM_, and T_EM_ cells in the spleen of recipient mice on day one (**A)** and day seven (**B**) after allo-HSCT. (**C** and **D**) Representative dot plots (left) and proportions (right) of CD8^+^ T_naïve_, T_CM_, and T_EM_ cells in the spleen of recipient mice on day one (**C**) and day seven (**D**) after allo-HSCT. Data are presented as means ± SEMs and analyzed by unpaired t test.

## DISCUSSION

In this study, through retrospective analysis of clinical data, we found that altering the timing of stem cell infusion can unexpectedly impact the incidence and severity of aGVHD in patients receiving ISD or HID-HSCT. Notably, patients received stem cell infusions before 2:00 pm had significantly less incidence and severity of aGVHD, lower TRM and better survival than did those treated after 2:00 pm. Currently, in medical practice, timing decisions for stem cell infusions typically hinge on staff convenience. Our results suggest that, pending validation through prospective multicenter studies that include varied graft types, prioritizing infusions before early afternoon could improve outcomes in allo-HSCT procedures. Making such scheduling arrangements holds promise for substantially improving patient prognosis and quality of life.

Our animal study corroborated that stem cell infusions administered early (ZT14) in the day significantly attenuate the risk of aGVHD following allo-HSCT compared to those administered late (ZT5) in the day. A key finding is that the recipient’s circadian rhythm plays a critical role in this dynamic. Mechanistically, compared with those in the late-infused group, the mice in the early-infused group exhibited significantly lower levels of the pro-inflammatory cytokine IL-1α following the conditioning regimen and subsequent reductions in T-cell activation and differentiation after transplantation. Furthermore, we observed significantly lower percentages of effector memory T cells in the ZT14 group than in the ZT5 group. These insights align with the current understanding that the incidence and severity of aGVHD are intricately associated with the levels of pro-inflammatory cytokines and T cell status(Blazar et al., 2012; Cooney et al., 1991; Welniak et al., 2000). The immune system is highly regulated by the circadian rhythm(Wang et al., 2022). In the future, exploring the mechanisms by which circadian timing influences the production of pro-inflammatory cytokines and T cell activation and differentiation in the context of allo-HSCT will be of substantial interest.

The often-neglected influence of circadian rhythms is emerging as a pivotal factor in clinical practice(Klerman et al., 2022; Levi et al., 1997; McKenna et al., 2018). For instance, a recent study of melanoma immunotherapy indicated that administering immune checkpoint inhibitors prior to 4:30 pm markedly improved treatment outcomes compared to later infusions(Qian et al., 2021). Our research bolsters the idea that administering the appropriate intervention at the optimal time can improve patient prognosis. To the best of our knowledge, this study is the first to identify the circadian timing of stem cell infusion as an independent risk factor for aGVHD development. Given the significant and far-reaching impact on aGVHD and long-term survival achieved by simply altering the timing of stem cell infusion, we believe that the timing of stem cell infusion itself may represent a critical window for clinical intervention. Our findings also highlight the importance of considering circadian factors when evaluating the effectiveness of drugs and treatments for aGVHD. Given that aGVHD following allo-HSCT is inherently a transplant-related immune rejection condition—a phenomenon prevalent in diverse organ transplant scenarios—it is of great interest to investigate how circadian rhythms may affect immune rejection in other types of organ transplantations.

## Data Availability

The full data set will be available on the Vivli platform with publication. This paper does not report original code. Any additional information required to reanalyze the data reported in this paper is available from the lead contact upon request.

https://vivli.org/

## ACKNOWLEDGMENTS

We thank Dr. Haiming Wei’s laboratory at the USTC for assisting in the development of the aGVHD mouse model. We thank the animal facility and flow cytometry facility at USTC for their technical support. We appreciate the insightful suggestions and comments provided by the Neuroscience Pioneer Club to improve this manuscript. Cheng Zhan is supported by grants from the Research Funds of Center for Advanced Interdisciplinary Science and Biomedicine of IHM (QYPY20220018), the National Natural Science Foundation of China (32271063, 31822026, 31500860), and the National Science and Technology Innovation 2030 Major Project of China (2021ZD0203900). Xiaoyu Zhu is supported by grants from the National Natural Science Foundation of China (82270223), the Anhui Provincial Department of Education Scientific Research Project (2023AH010079) and the Anhui Provincial Natural Science Foundation (2308085J09).

## AUTHOR CONTRIBUTIONS

Cheng Zhan and Xiaoyu Zhu conceived the study and wrote the manuscript. Yue Wu, Baolin Tang, Kaidi Song, and Guangyu Sun contributed to the clinical data collection and data analysis. Yiwen Hou, Zhonglin Zhang and Zhiwei Liu developed the aGVHD mouse model and evaluated the pathological score. Yiwen Hou and Liang Wang performed the flow cytometry experiments. The data analysis presented in this study was carried out by Yue Wu and Yiwen Hou. Yue Wu and Yiwen Hou contributed equally to this manuscript.

## Supplementary Figures and Tables

**Figure S1.**
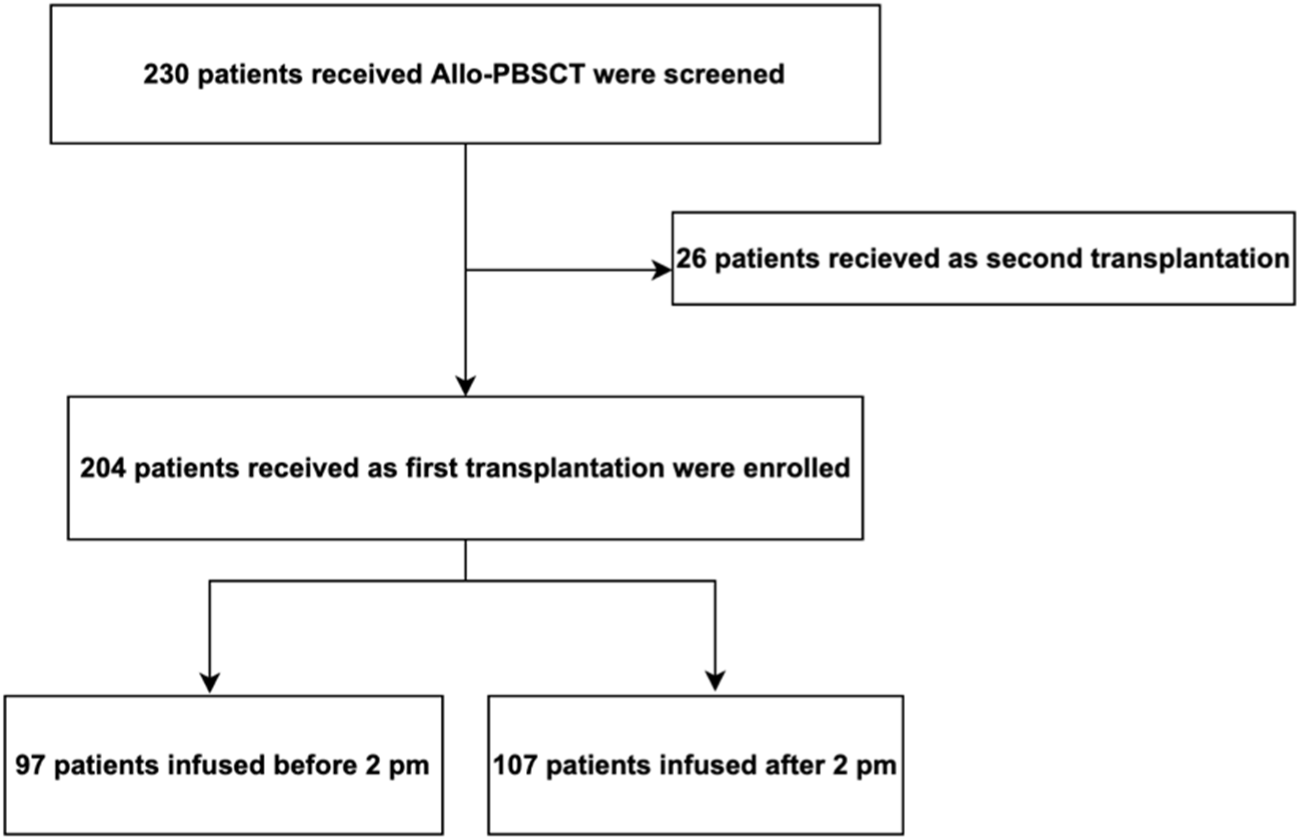
Diagram of patients enrolled in this study. Allo-PBSCT allogeneic peripheral blood stem cell transplantation.

**Figure S2.**
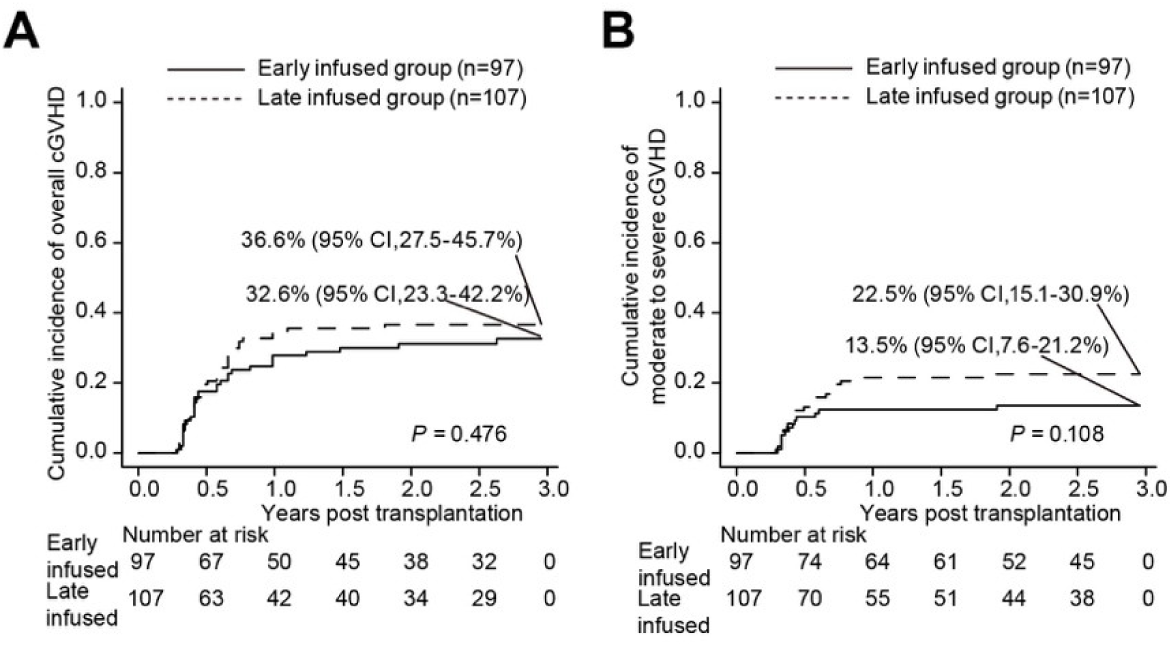
Cumulative incidences of overall cGVHD (A) and moderate to severe cGVHD (B) in the early infused group and late infused group.

**Figure S3.**
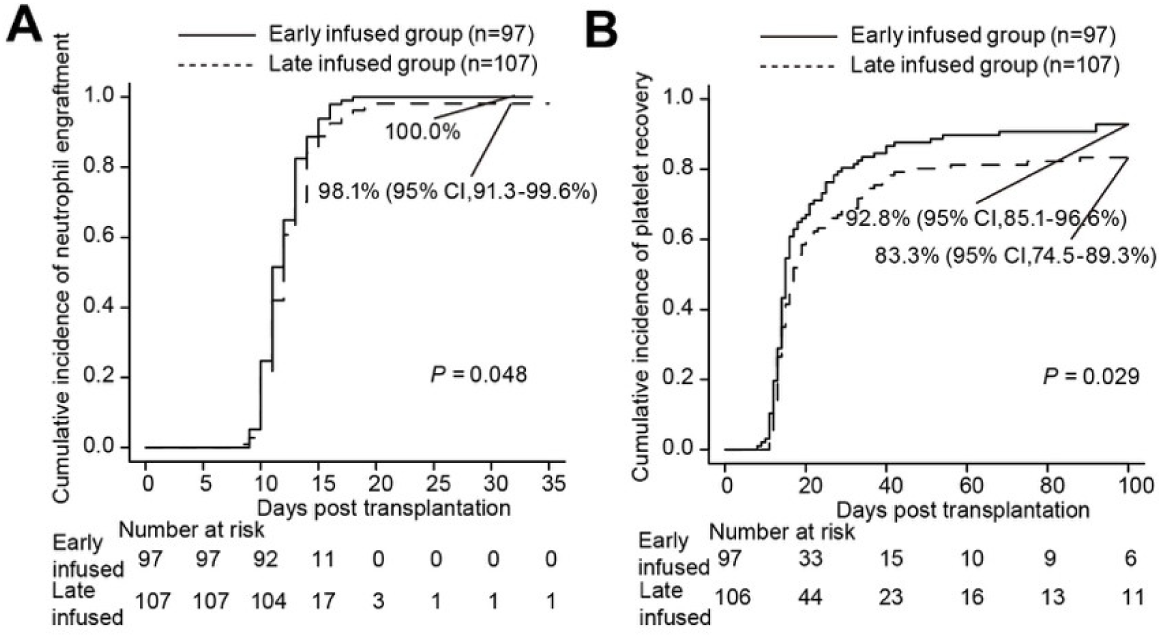
Cumulative incidences of neutrophil engraftment by day 42 (A) and platelet recovery by day 100 (B) in the early infused group and late infused group.

**Figure S4.**
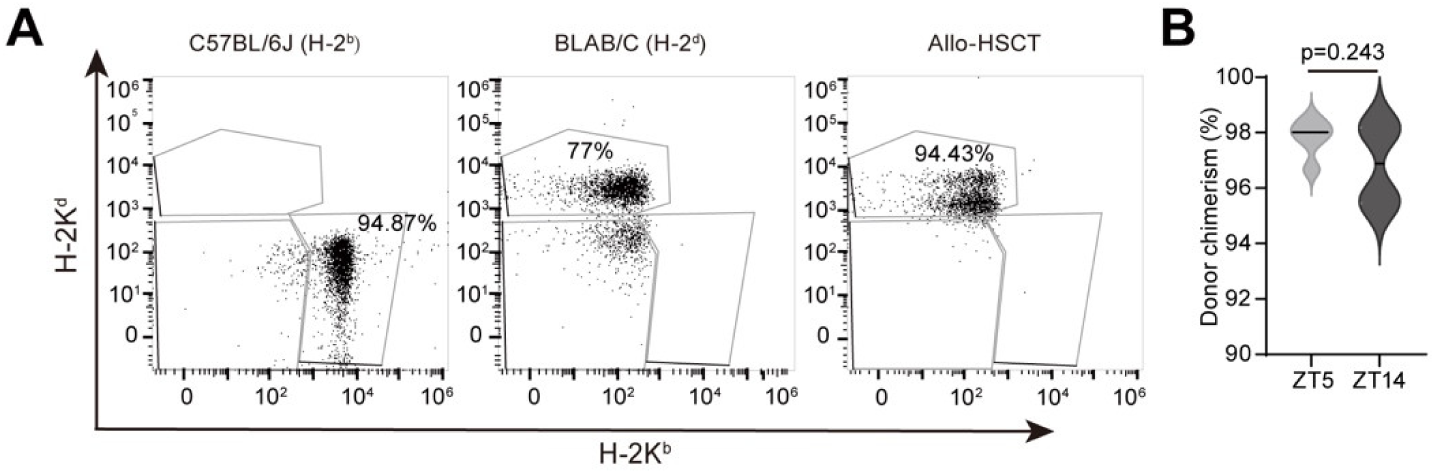
Impact of stem cell infusion timing on donor chimerism. (**A**) H-2K^b^ and H-2K^d^ staining of peripheral blood in untransplanted C57BL/6J mice (left), BALB/c mice (middle), and transplanted C57BL/6J mice (right) on day14 following allo-HSCT. (**B**) Donor chimerism in peripheral blood of ZT5 and ZT14 groups on day 14 after allo-HSCT (ZT5 n=5, ZT14 n=4). Data are presented as means ± SEM and analyzed using unpaired *t* test.

**Figure S5.**
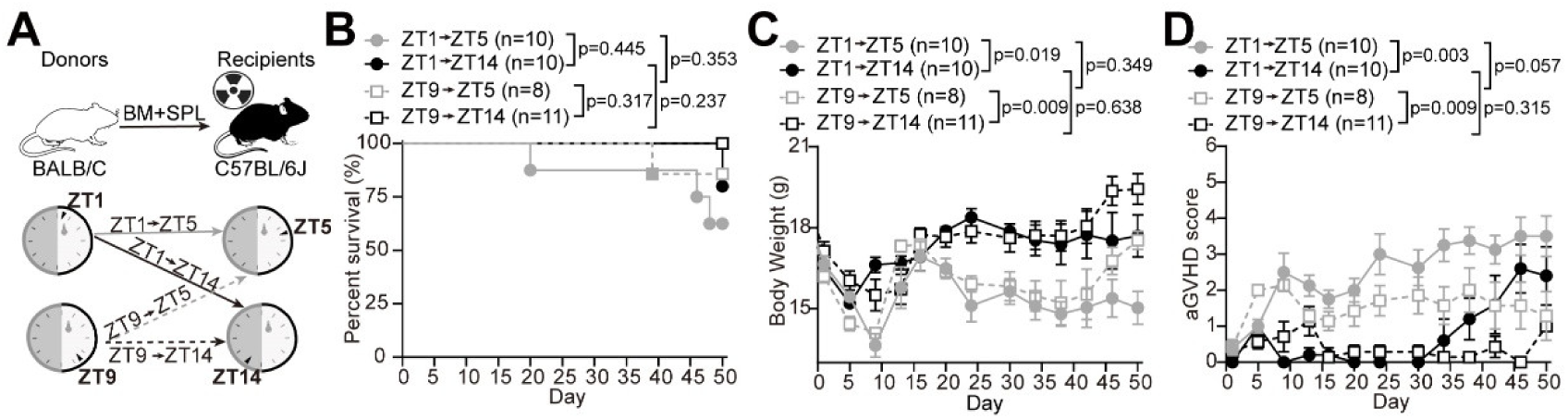
Impacts of the rhythms of recipient and donor mice on aGVHD development. (**A**) Experimental schematic. Both ZT5 and ZT14 recipient mice received BM cells and splenocytes transplantation from ZT1 or ZT9 donor mice. (**B-D**) Survival rate (**B**), body weight (**C**) and aGVHD score (**D**) of recipient mice. Data are presented as means ± SEM and analyzed by two-way ANOVA followed by Bonferroni post hoc test (**C** and **D**) and log-rank test (**B**).

**Figure S6.**
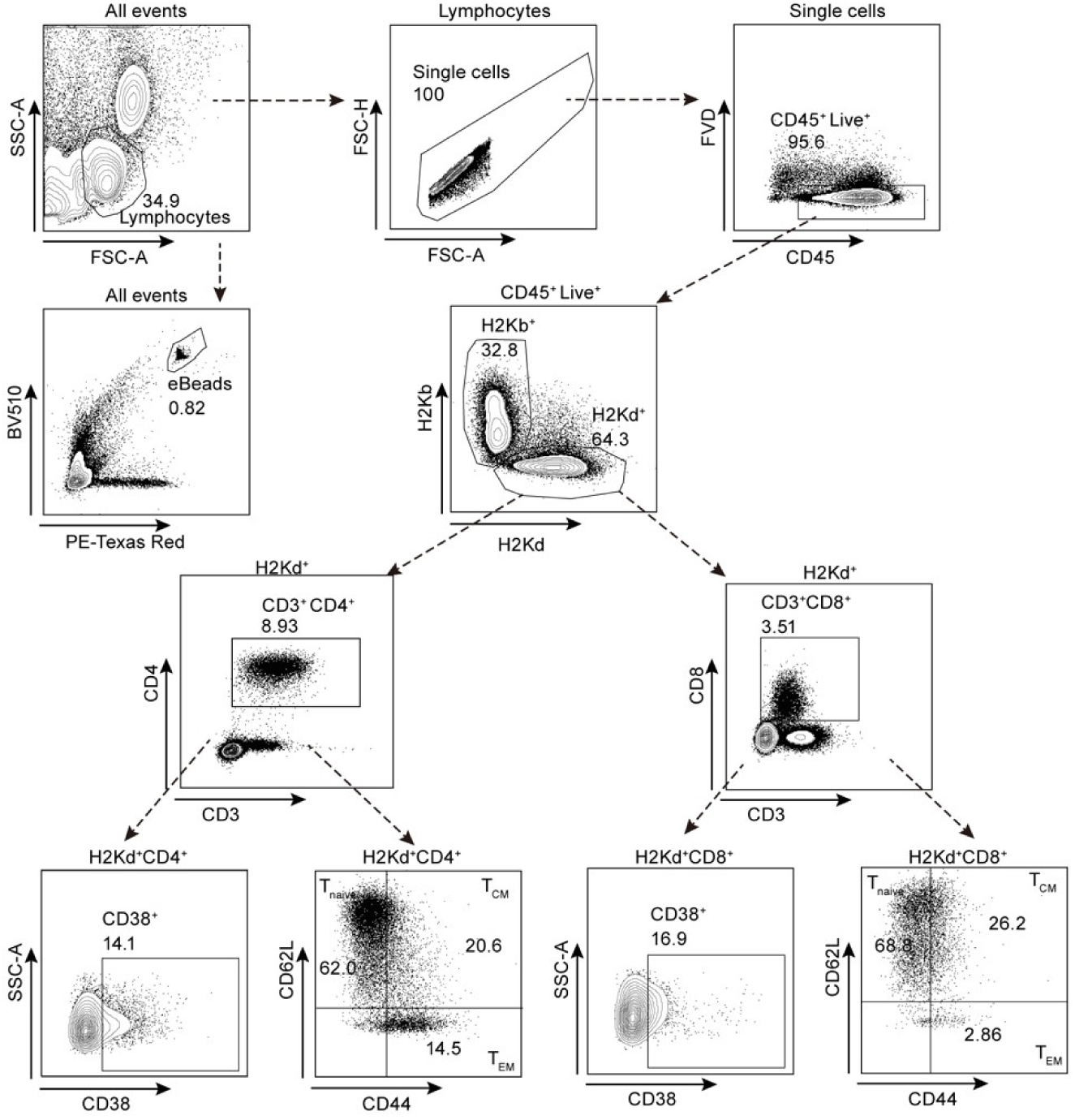
Gating strategy in flow cytometry analysis. Starting from upper left panel and going down following the arrows in dotted lines, we gated successively: lymphocytes, CD45^+^ live cells, H2kd^+^ donor cells, and T (CD3^+^) cells. Donor T cells were subdivided into CD4^+^ T and CD8^+^ T-cell subsets. Naïve T cells (CD44^−^CD62L^+^, T_naïve_), central memory T cells (CD44^+^CD62L^+^, T_CM_), and effector memory T cells (CD44^+^CD62L^−^, T_EM_) were gated according to the expression of CD44 and CD62L.CounteBeads were added to cell suspensions for absolute cell counting.

**Table S1.** Characteristics of patients between the early infused group and late infused group.

| Characteristics | early infused group<br>(n=97) | late infused group<br>(n=107) | *P* |
| --- | --- | --- | --- |
| Sex, n (%) |  |  | 0.780 |
| Male | 53 (54.6) | 56 (52.3) |  |
| Female | 44 (45.4) | 51 (47.7) |  |
| Age (years), Median (Range) | 32 (4-61) | 32 (3-62) | 0.279 |
| Weight(kg), Median (Range) | 60 (14-92) | 62 (14-109) | 0.967 |
| Diagnostics, n (%) |  |  | 0.385 |
| AML | 42 (43.3) | 36 (33.6) |  |
| ALL | 19 (19.6) | 17 (15.9) |  |
| MDS/MPN | 19 (19.6) | 32 (29.9) |  |
| AA | 15 (15.5) | 20 (18.7) |  |
| Others* | 2 (2.1) | 2 (1.9) |  |
| Donor type, n (%) |  |  | 1.000 |
| ISD | 65 (67.0) | 72 (67.3) |  |
| HID | 32 (33.0) | 35 (32.7) |  |
| CD34<sup>+</sup>cell infused (10<sup>6</sup>/kg) | 5.62 (0.32-67.5) | 4.90 (0.66-36.30) | 0.375 |
| TNCs infused (10<sup>8</sup>/kg) | 9.95 (2.50-101.32) | 9.59 (2.35-56.95) | 0.284 |
| Female to male, n (%) |  |  | 0.333 |
| Yes | 21 (21.6) | 30 (28.0) |  |
| No | 76 (78.4) | 77 (72.0) |  |
| Conditioning regimen, n (%) |  |  | 1.000 |
| MAC | 74 (76.3) | 82 (76.6) |  |
| RIC | 23 (23.7) | 25 (23.4) |  |
| MTX prophylaxis, n (%) | 8 (8.2) | 12 (11.2) | 0.638 |
| Prophylaxis drug, n (%) |  |  | 0.991 |
| Without rATG or PT-Cy | 65 (67.0) | 72 (67.3) |  |
| With rATG only | 10 (10.3) | 12 (11.2) |  |
| With PT-Cy only | 14 (14.4) | 14 (13.1) |  |
| With both rATG and PT-Cy | 8 (8.2) | 9 (8.4) |  |
| ABO incompatibility, n (%) |  |  | 0.844 |
| Identical | 51 (52.6) | 60 (56.1) |  |
| Minor incompatibility | 19 (19.6) | 24 (22.4) |  |
| Major incompatibility | 16 (16.5) | 18 (16.8) |  |
| Bidirectional incompatibility | 11 (11.3) | 5 (4.7) |  |
| Follow-up time among survivors (month) | 50.3 (7.1-106.1) | 42.2 (7.5-101.9) | 0.284 |

ISD identical sibling donor, HID haplo-identical donor, TNCs total nucleated cells, MAC myeloablative conditioning regimen, RIC reduced intensity conditioning, MTX methotrexate, rATG rabbit anti-thymocyte globulin, PT-Cy post-transplant cyclophosphamide, \*MPAL, mixed phenotype acute leukemia; NHL, non-Hodgkin's lymphoma

**Table S2.**
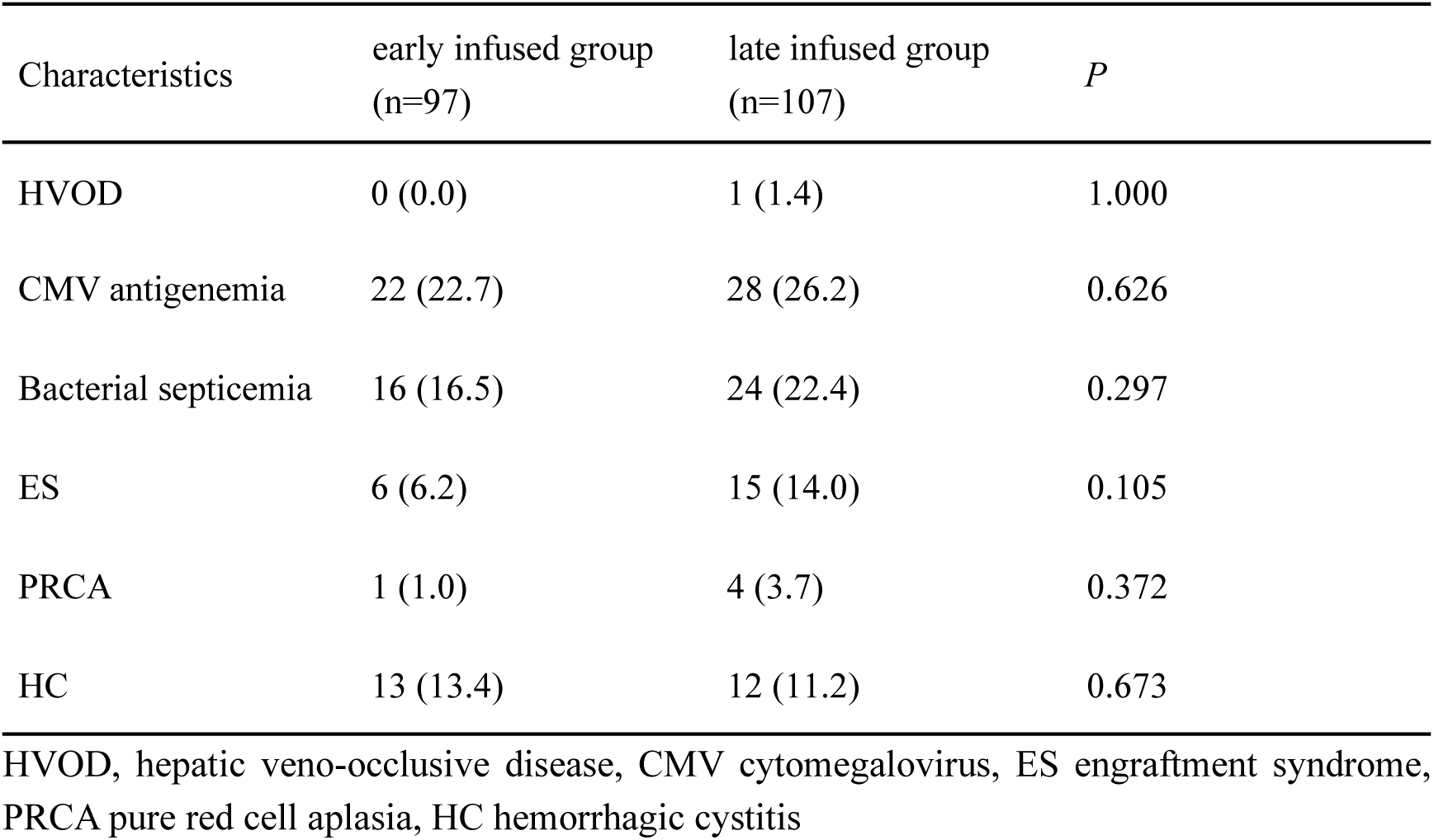
Complications post transplantation between the early infused group and late infused group.

**Table S3.** Causes of death of the early infused group and late infused group.

| Cause | early infused group | late infused group |
| --- | --- | --- |
| Relapse | 12 (52.2) | 11 (29.7) |
| Uncontrolled acute GVHD | 2 (8.7) | 7 (18.9) |
| Extensive chronic GVHD | 1 (4.3) | 3 (8.1) |
| Infection | 3 (13.0) | 8 (21.6) |
| MODS | 4 (17.4) | 6 (16.2) |
| Others | 1 (4.3) | 2 (5.4) |

GVHD graft-versus-host disease, MODS, multi-organ dysfunction syndrome

## Key resources table

| REAGENT or RESOURCE | SOURCE | IDENTIFIER |
| --- | --- | --- |
| **Antibodies** |  |  |
| InVivoMAb anti-mouse CD16/CD32(2.4G2) | BioXcell | BE0307 |
| APC-CY7-anti mouse CD45 Monoclonal Antibody(30-F11) | BioLegend | 103116 |
| PE-Cy5-anti mouse CD3 Monoclonal Antibody (17A2) | eBioscience | 15-0032-82 |
| FITC-anti mouse CD4 Monoclonal Antibody(RM4-5) | eBioscience | 11-0042-82 |
| AF700-anti mouse CD8A Monoclonal Antibody(53-6.7) | eBioscience | 56-0081-82 |
| Super Bright 780 anti-CD44 | eBioscience | 78-0441-82 |
| super bright 600 anti mouse CD62L (L-Selectin) Monoclonal Antibody (MEL-14) | ebioscience | 63-0621-82 |
| PE-Cyanine7 anti mouse CD38 Monoclonal Antibody (90) | ebioscience | 25-0381-82 |
| APC anti-mouse H-2Kd | Biolegend | 116620 |
| PE anti-mouse H-2Kb | Biolegend | 116507 |
| Fixable Viability Dye eFluor™ 506 | eBioscience | 65-0866-18 |
| **Biological samples** |  |  |
| Patient-derived allo-HSCT | The First Affiliated Hospital of the University of Science and Technology of China | N/A |
| **Chemicals, peptides, and recombinant proteins** |  |  |
| Gentamicin Sulfate | BioSharp | BS143-5g |
| RPMI 1640 medium | Gibco | C22400500BT |
| Fetal bovine serum | Biological Industries | C04001-500 |
| RBC Lysing Buffer | BD Pharm Lyse | 555899 |
| BSA | Thermo Fisher Scientific | No. 15260037 |
| CounteBeads | eBioscience | 01-1234-42 |
| **Critical commercial assays** |  |  |
| Cytometric Bead Array (CBA) mouse cytokine kit | BD Biosciences | 560485 |
| **Experimental models: Organisms/strains** |  |  |
| Mouse: C57BL/6J | GemPharmatech Co., Ltd. | N000013 |
| Mouse: BALB/c | Shanghai Silaike Experiment Animal Co., Ltd. | N/A |
| **Software and algorithms** |  |  |
| FlowJo v10 software | FlowJo | https://flowjo.becton dickinson.cn |
| FCAP Array Software | BD Biosciences | https://legendplex.qognit.com/workflow |
| R software packages | R Foundation for Statistical Computing | https://www.r-project.org |
| GraphPad Prism 9.0 | GraphPad Software | https://www.graphpad.com |

## STAR★METHODS

### RESOURCE AVAILABILITY

#### Lead contact

Further information and requests for resources and reagents should be directed to and will be fulfilled by the lead contact, Cheng Zhan.

#### Materials availability

This study did not generate any new unique reagents.

#### Data and code availability

The full dataset will be available on the Vivli platform (https://vivli.org/) with publication. This paper does not report any original code. Any additional information required to reanalyze the data reported in this paper is available from the lead contact upon request.

## EXPERIMENTAL MODEL AND STUDY PARTICIPANT DETAILS

### Patient and transplantation characteristics

All patients received mobilized peripheral blood stem cells as a graft source. The study included 169 patients with malignant hematologic diseases and 35 patients with aplastic anemia (AA), with the most frequent diagnosis of hematological malignances being acute leukemia (55.9%). The median age of the whole cohort was 32 (range, 3-62) years, and 109 (53.4%) participants were male. The median infused total nucleated cell (TNC) count was 9.73 (range, 2.35 to 101.32) ×10^8^/kg, and the median infused CD34^+^ cell count was 5.12 (range, 0.32 to 67.5) ×10^6^/kg. Among all the patients, in addition to cyclosporine (CsA) and mycophenolate mofetil (MMF), 22 patients received rabbit anti-thymocyte globulin (rATG), 28 patients received post-transplant cyclophosphamide (PT-Cy), 17 patients received both rATG and PT-Cy, and 20 patients received short-term methotrexate (MTX) for GVHD prophylaxis. All surviving patients were followed until 31 Dec 2023, and the median follow-up time was 47.5 (range, 7.1 to 106.1) months.

The protocol was approved by the Ethics Committee of the First Affiliated Hospital of the University of Science and Technology of China (USTC) (REC reference: 2024/RE/49). The patients or guardians provided informed consent before transplantation for the use of their data for research, in accordance with the Declaration of Helsinki.

### Mice

C57BL/6J (H2k^b^) and BALB/c (H2k^d^) mice were purchased from GemPharmatech Co., Ltd. (China) and Shanghai Silaike Experiment Animal Co., Ltd., respectively. The animals were maintained in a specific pathogen-free animal facility for at least one week before any further experiments were conducted. All mice were group-housed under a stable environment (22-25°C ambient temperature, humidity 40%-70%) with a 12 h light/12 h dark cycle (lights on from 8 am to 8 pm) and *ad libitum* access to water and a standard chow diet (SPF Rodent Feed; 4% fat, 18% protein, 5% crude Fiber; WQJX BIO-Technology, China). All the experiments were performed on adult mice (9-17 weeks old). The care and use of animals in this study were approved by the Institutional Animal Use and Care Committee of the University of Science and Technology of China (USTC) (USTCACUC26110122040) and conformed to the institutional guidelines of the USTC, as well as governmental regulations.

## METHOD DETAILS

### HLA typing, stem cell mobilization and collection

HLA typing of donors and patients was performed using real-time PCR with a high resolution of 10 alleles for HLA-A, HLA-B, HLA-C, HLA-DRB1, and HLA-DQB1. The graft source was unmanipulated mobilized PBSCs in all patients. The donors received four consecutive days of granulocyte colony-stimulating factor (G-CSF) (filgrastim) (10 mg/kg/d) for peripheral stem cell mobilization followed by apheresis on day 5. The target stem cell doses for collection and infusion for patients were 5 × 10^8^ total nucleated cells (TNCs)/kg (of recipient weight) and 2 × 10^6^ CD34^+^ cells/kg, respectively. Once the stem cells were collected, they were immediately infused into the patient through the central venous catheter.

### Conditioning regimen and GVHD prophylaxis

A total of 156 of 204 (76.5%) patients received myeloablative conditioning (MAC), and 48 patients (23.5%) received a reduced-intensity conditioning (RIC) regimen. RIC conditioning regimens were performed in patients who were either older than 40 years or had comorbid severe infection before transplantation. For MAC, 146 patients received a busulfan (Bu)/cyclophosphamide (Cy)-based regimen (Bu, 0.8 mg/kg every 6 hours for 3 or 4 days; Cy, 60 mg/kg daily for 2 days) supplemented with fludarabine (Flu) (30 mg/m^2^ daily for 4 days), and 10 patients received a fractionated total body irradiation (TBI) or total marrow irradiation (TMI)/Cy regimen (TBI, total 12 Gy, 4 fractions or TMI total 15 Gy, 3 fractions; Cy, 60 mg/kg daily for 2 days), of which 5 patients with cytarabine (Ara-C) added (2 g/m^2^ daily for 4 days). For RIC, 35 patients received an irradiation-based conditioning regimen consisting of Flu 30-50 mg/m^2^ for 4-5 days, Cy 14.5-60 mg/kg daily for 2 days and TBI/TMI total 3-4 Gy, 13 patients received a Bu-based conditioning regimen consisting of Flu 30-50 mg/m^2^ for 4-5 days, Cy 60 mg/kg daily for 2 days and Bu 0.8 mg/kg every 6 hours for 2 days.

To prevent GVHD, all patients were given a combination of cyclosporine (CsA) and mycophenolate mofetil (MMF). Twenty-two (10.8%) patients received additional rabbit antithymocyte globulin (rATG), 28 (13.7%) patients received additional post-transplant cyclophosphamide (PT-Cy), 17 (8.3%) patients received both rATG and PT-Cy, and 20 (9.8%) patients were given extra short-term methotrexate (MTX) for GVHD prevention. CsA was started intravenously (2.5-3 mg/kg/day) on day −1 with a target level of 250-300 ng/mL for at least one month before switching to oral intake, and this concentration was tapered by day +60 based on the presence of GVHD or disease conditions to maintain the trough concentration at 100-150 ng/mL and withdrawn by +100 in the absence of GVHD. MMF (25-30 mg/kg/day, p.o. three times per day) was started on day +1 and continued until day +35. rATG (thymoglobulin) was administered from day −6 to −4 at a total dose of 4.5-7.5 mg/kg. PT-Cy was administered on days +3 and +4 at a total dose of 100 mg/kg. MTX was administered intravenously on day +1 at 15 mg/m^2^ and 10 mg/m^2^ on days +3 and +6 post-transplantation. The other supportive treatments adhered to standard protocols based on previously published criteria^1^.

### Definitions

Neutrophil recovery was defined as the first of three consecutive days with a neutrophil count of at least 0.5×10^9^/L, and primary platelet recovery was defined as the first day with a platelet count of at least 50×10^9^/L for seven consecutive days without transfusion support. Primary graft failure was defined as failure to achieve an absolute neutrophil count (ANC) of 0.5×10^9^/L with the absence of donor hematopoiesis by 30 days after transplantation. aGVHD and cGVHD were diagnosed and classified using published criteria^2,3^. Disease recurrence was diagnosed using clinical and pathological standards. Overall survival (OS) was defined as the time from the first day of transplantation until death from any cause or the last follow-up date. Disease-free survival (DFS) was defined as the interval between transplantation and disease recurrence, death or the last follow-up date, whichever occurred first. The composite endpoint of GVHD-free, relapse-free survival (GRFS) was defined as the first event occurring within 36 months after transplantation among Grade III to IV aGVHD, moderate to severe cGVHD, relapse, or death for any reason.

### Induction of aGVHD in mice

Major histocompatibility complex (MHC)-mismatched transplants were performed using BALB/c and C57BL/6J mice as donors and recipients, respectively. Female C57BL/6J mice (9 weeks old) received a single fraction of 1100 cGy (^137^Cs source) irradiation 5-6 hours prior to transplantation and were intravenously administered 1×10^7^ bone marrow cells plus 6×10^7^ splenocytes from male BALB/c mice (9 weeks old). Starting 2 days before irradiation and continuing until 1 week after transplantation, gentamicin (320 mg/L) was added to the drinking water of the recipients.

### Donor cell collection

For the induction of aGVHD in mice, bone marrow cells were harvested from donor femurs and tibias and resuspended in RPMI 1640 medium (C22400500BT, Gibco) supplemented with 2% fetal bovine serum (FBS) (C04001-500, Biological Industries, Israel). Splenocytes were obtained by gently crushing the spleen through 40 μm mesh filters in RPMI 1640 medium supplemented with 2% FBS to release the cells. Erythrocytes were lysed by incubating the cells in RBC Lysing Buffer (555899, BD Pharm Lyse) for 3 min at room temperature. The preparations were then filtered through a 40 μm strainer to remove debris and washed twice in DPBS. The cells were then counted using a cell counter (Counter star, China) and resuspended in 0.2 mL of DPBS for injection.

### Flow cytometry

To evaluate the activation and differentiation of T cells, the spleen was isolated and processed through a 40-μm strainer in RPMI 1640 medium supplemented with 20 mg/ml BSA (Thermo Fisher Scientific). After blocking with an anti-mouse CD16/CD32 mAb (1 μg/ ml, BE0307, BioXcell) for 10 min at 4°C to prevent nonspecific antibody binding, the cell suspension was co-incubated with APC-Cy7 anti-CD45 (1 μg/ ml, 30-F11), PE-Cy5 anti-CD3 (1 μg/ ml, 17A2), FITC ant-CD4 (1 μg/ ml, RM4-5), AF700 anti-CD8 (1 μg/ ml, 53-6.7), Super Bright 780 anti-CD44 (1 μg/ ml, IM7), Super Bright 600 anti-CD62L (L-selectin) (1 μg/ ml, MEL-14), PE-Cy7 anti-CD38 (1 μg/ ml, 90), APC anti-H-2K^d^ (1 μg/ ml, SF1-1.1) and PE anti-H-2Kb (1 μg/ ml, AF6-88.5) for 30 min at 4 ℃. Fixable viability dye (FVD, eFluor 506, eBioscience) staining was used to exclude dead cells from viable cells. After erythrocytes were lysed with RBC Lysis Buffer and washed once in fluorescence-activated cell sorting (FACS) buffer solution, the cells were then passed through a 40 μm cell strainer. The cells were resuspended in 200 μl of FACS buffer solution and analyzed by a FACSAria II (BD Biosciences) and FlowJo v10 software (FlowJo, Ashland, OR, USA). A total of 20 μl of microparticles per sample (CounteBeads, 01-1234-42, eBioscience) was added to the cell suspensions for absolute counting by flow cytometry. The following equation was used to calculate the absolute cell number (cells / sample): (cell count × eBeads volume × eBeads concentration) / eBeads count.

For evaluation of peripheral chimerism, blood was first incubated with purified anti-mouse CD16/CD32 mAb for 10 min at 4 ℃ and then co-incubated with APC anti-H-2K^d^ (1 μg/ ml) and PE anti-H-2K^b^ (1 μg/ ml) for 30 min at 4 ℃. After staining with fixable viability dye, the cell suspensions were analyzed with a Beckman Coulter Cytoflex (Beckman Coulter, Brea, CA, USA). The obtained data were analyzed using FlowJo v10 software (FlowJo, Ashland, OR, USA). PE-H-2K^b^ and APC-H-2K^d^ single staining was performed on blood from naive C57BL/6J and BALB/c mice as positive controls to establish compensatory parameters. Unstained cells were used to establish the analysis area. The percentages of H-2K^b+^ and H-2K^d+^ cell subpopulations were calculated. The donor chimerism was calculated according to the equation: Donor chimerism (%) = H-2k^d^ % / (H-2K^b^ %+H-2K^d^ %) ×100%.

### aGVHD scoring

Following transplantation, the recipients were monitored for survival once daily and for the occurrence of aGVHD symptomatology twice weekly. The severity of aGVHD was assessed by scoring weight loss, activity, posture (hunching), and fur-ruffling. Each criterion was scored on a scale of 0 to 2, with 0 representing normal, 1 representing mildly abnormal, and 2 representing severely abnormal. The aGVHD clinical scores were calculated by summing the four criteria scores. Mice that were moribund or scored above 5 were euthanized.

### Measurement of cytokine levels

Blood was drawn from the retrobulbar intraorbital capillary plexus of the recipients and collected in sterile tubes. A Cytometric Bead Array (CBA) mouse cytokine kit (560485, BD Biosciences) was used to measure the serum cytokine levels according to the manufacturer’s instructions. FCAP Array Software (v3.0, 652099, BD Biosciences) was used for CBA data analysis.

### Histology and imaging

After an overdose of the anesthetic 2,2,2-tribromoethanol (> 400 mg/kg, i.p.) was injected, the animals were cardiac-perfused with cold phosphate-buffered saline (PBS), followed by 4% paraformaldehyde. The skin, liver, colon, and lungs were harvested and then postfixed in 4% paraformaldehyde overnight with cryoprotection in 30% sucrose. The tissue sections (5 µm) were cut with a freezing cryostat (Leica CR 1900) and stained with hematoxylin and eosin. Montage images were acquired and stitched using an automated slider scanner (Exo, Meca Scientific, China). The brightness, contrast, and pseudocolor were adjusted using SlideViewer (Meca Scientific, China).

### Pathologic tissue scoring

Histopathological assessments of the skin, liver, colon, and lungs were performed as previously described^4,5^. Skin clinical scores were calculated (0 to 10 grades) as the sum of scores for the following criteria: epidermis (0, normal; 1, foci of interface damage in <20% of sections with occasional necrotic keratinocytes; 2, widespread interface damage in >20% of sections); dermis (0, normal; 1, slightly altered with mildly increased collagen density; 2, markedly increased collagen density); inflammation (0, none; 1, focal infiltrates; 2, widespread infiltrates); s.c., fat (0, normal; 1, reduced number of normal adipocytes; 2, serous fat atrophy); and follicles (0, normal number of hair follicles, ∼5 per linear millimeter; 1, between 1 and 5 follicles per linear millimeter; 2, <1 follicle per linear millimeter).

Liver sections were scored (0 to 4 grades) based on bile duct injury (manifest by nuclear hyperchromasia, nuclear crowding, infiltrating lymphocytes, and cytoplasmic vacuolation) and inflammation (infiltration with lymphocytes, neutrophils, and eosinophils) according to the following criteria: the number of involved tracts and the severity of disease in each tract (0, none; 1, few involved tracts with mild involvement; 2, numerous involved tracts but with only mild disease; 3, injury in the majority of tracts; and 4, severe involvement of most tracts).

Colon sections were scored (0 to 4 grades) based on crypt apoptosis (0, rare to none; 1, occasional apoptotic bodies per 10 crypts; 2, few apoptotic bodies per 10 crypts; 3, the majority of crypts contain an apoptotic body; 4, the majority of crypts contain >1 apoptotic body) and inflammation (0, none; 1, mild; 2, moderate; 3, severe, without ulceration; 4, severe, with ulceration).

Lung sections were scored (0 to 4 grades) based on perivascular and peribronchiolar cuffing and infiltration for the following criteria: 0, normal; 0.5, minimal perivascular cuffing; 1, perivascular cuffing; 1 to 2 cells in thickness, involving up to 15% of vessels; 1.5, perivascular cuffing; 1 to 2 cells in thickness, involving up to 15% of vessels and infiltration into parenchyma proper; 2, perivascular cuffing; 2 to 3 cells in thickness, involving up to 25% of vessels and infiltration into parenchyma proper; 2.5, perivascular cuffing; 2 to 3 cells in thickness, involving 25% to 50% of vessels and infiltration into parenchyma proper; 3, perivascular cuffing; 4 to 5 cells in thickness, involving 25% to 50% of vessels, peribronchiolar cuffing (2 to 3 cells) and infiltration into parenchyma proper; 3.5, perivascular cuffing; 6 to 7 cells in thickness, involving greater than 50% of vessels, peribronchiolar cuffing (4 to 5 cells); and 4, perivascular cuffing, greater than 8 cells in thickness, involving greater than 50% of vessels, peribronchiolar cuffing (>6 cells).

## QUANTIFICATION AND STATISTICAL ANALYSIS

### Statistical analysis of clinical data

The quantitative variables are represented by the median and range. The qualitative factors are expressed as frequencies and proportions. Quantitative variables were calculated and compared using Student’s *t* test or the Mann‒Whitney U test. Qualitative variables were analyzed using the chi-square test or Fisher’s exact test. Ordinal data were analyzed using the Cochran-Armitage trend test. To calculate the cumulative incidence of engraftment, aGVHD, cGVHD, transplant-related mortality (TRM), and relapse, cumulative incidence curves were constructed in a competing risk setting, with death considered a competing event, and the results were compared using Gray’s test ^15^. TRM and relapse were regarded as competing risk factors. The Kaplan‒Meier method and log-rank test were used to compare the probabilities of 3-year overall survival (OS), disease-free survival (DFS), and GVHD free, relapse-free survival (GRFS).

To determine the risk factors for Grade II-IV and Grade III-IV aGVHD, we conducted univariate and multivariate analyses with a Fine-Gray proportional hazards regression model^16^. In the regression model, we included the timing of stem cell infusion as an ordinal variable in the univariate and multivariate analyses. All variables with a 2-sided *P* value less than 0.2 were included in the multivariate analysis. The R software package, version 4.2.2 (R Foundation for Statistical Computing, Vienna, Austria), was used for statistical analyses. A 2-sided *P* value less than 0.05 was considered to indicate statistical significance.

### Statistical analysis of the mouse experiments

Statistical analysis was performed using GraphPad Prism 9.0 (GraphPad Software, CA, USA). Statistical differences in the data were determined using an unpaired Student’s *t* test for two-group comparisons, two-way analysis of variance (ANOVA) followed by the Bonferroni post hoc test for multiple comparisons, and the log-rank test for survival (Kaplan–Meier survival curves) difference comparisons, as indicated in all the figure legends. The data are shown as the means ± SEMs.

## REFERENCES

Aardal, N.P., and Laerum, O.D. (1983). Circadian variations in mouse bone marrow. Exp Hematol 11, 792–801.

Abrahamsen, J.F., Smaaland, R., Sothern, R.B., and Laerum, O.D. (1998). Variation in cell yield and proliferative activity of positive selected human CD34+ bone marrow cells along the circadian time scale. Eur J Haematol 60, 7–15.

Bacigalupo, A., Lamparelli, T., Bruzzi, P., Guidi, S., Alessandrino, P.E., di Bartolomeo, P., Oneto, R., Bruno, B., Barbanti, M., Sacchi, N., et al. (2001). Antithymocyte globulin for graft-versus-host disease prophylaxis in transplants from unrelated donors: 2 randomized studies from Gruppo Italiano Trapianti Midollo Osseo (GITMO). Blood 98, 2942–2947.

Blazar, B.R., Murphy, W.J., and Abedi, M. (2012). Advances in graft-versus-host disease biology and therapy. Nat Rev Immunol 12, 443–458.

Bolanos-Meade, J., Hamadani, M., Wu, J., Al Malki, M.M., Martens, M.J., Runaas, L., Elmariah, H., Rezvani, A.R., Gooptu, M., Larkin, K.T., et al. (2023). Post-Transplantation Cyclophosphamide-Based Graft-versus-Host Disease Prophylaxis. N Engl J Med 388, 2338–2348.

Cooney, R., Owens, E., Jurasinski, C., Gray, K., Vannice, J., and Vary, T. (1991). Interleukin-1 receptor antagonist prevents sepsis-induced inhibition of protein synthesis. Am J Physiol 267, E636–641.

D’Hondt, L., McAuliffe, C., Damon, J., Reilly, J., Carlson, J., Dooner, M., Colvin, G., Lambert, J.F., Hsieh, C.C., Habibian, H., et al. (2004). Circadian variations of bone marrow engraftability. J Cell Physiol 200, 63–70.

Ferrara, J.L., Abhyankar, S., and Gilliland, D.G. (1993). Cytokine storm of graft-versus-host disease: a critical effector role for interleukin-1. Transplant Proc 25, 1216–1217.

Ferrara, J.L., Levine, J.E., Reddy, P., and Holler, E. (2009). Graft-versus-host disease. Lancet 373, 1550–1561.

Flowers, M.E., Inamoto, Y., Carpenter, P.A., Lee, S.J., Kiem, H.P., Petersdorf, E.W., Pereira, S.E., Nash, R.A., Mielcarek, M., Fero, M.L., et al. (2011). Comparative analysis of risk factors for acute graft-versus-host disease and for chronic graft-versus-host disease according to National Institutes of Health consensus criteria. Blood 117, 3214–3219.

Jagasia, M., Arora, M., Flowers, M.E., Chao, N.J., McCarthy, P.L., Cutler, C.S., Urbano-Ispizua, A., Pavletic, S.Z., Haagenson, M.D., Zhang, M.J., et al. (2012). Risk factors for acute GVHD and survival after hematopoietic cell transplantation. Blood 119, 296–307.

Klerman, E.B., Brager, A., Carskadon, M.A., Depner, C.M., Foster, R., Goel, N., Harrington, M., Holloway, P.M., Knauert, M.P., LeBourgeois, M.K., et al. (2022). Keeping an eye on circadian time in clinical research and medicine. Clin Transl Med 12, e1131.

Levi, F., Zidani, R., and Misset, J.L. (1997). Randomised multicentre trial of chronotherapy with oxaliplatin, fluorouracil, and folinic acid in metastatic colorectal cancer. International Organization for Cancer Chronotherapy. Lancet 350, 681–686.

Lucas, D., Battista, M., Shi, P.A., Isola, L., and Frenette, P.S. (2008). Mobilized hematopoietic stem cell yield depends on species-specific circadian timing. Cell Stem Cell 3, 364–366.

Malik, A., and Kanneganti, T.D. (2018). Function and regulation of IL-1alpha in inflammatory diseases and cancer. Immunol Rev 281, 124–137.

McKenna, H., van der Horst, G.T.J., Reiss, I., and Martin, D. (2018). Clinical chronobiology: a timely consideration in critical care medicine. Crit Care 22, 124.

Mendez-Ferrer, S., Lucas, D., Battista, M., and Frenette, P.S. (2008). Haematopoietic stem cell release is regulated by circadian oscillations. Nature 452, 442–447.

Qian, D.C., Kleber, T., Brammer, B., Xu, K.M., Switchenko, J.M., Janopaul-Naylor, J.R., Zhong, J., Yushak, M.L., Harvey, R.D., Paulos, C.M., et al. (2021). Effect of immunotherapy time-of-day infusion on overall survival among patients with advanced melanoma in the USA (MEMOIR): a propensity score-matched analysis of a single-centre, longitudinal study. Lancet Oncol 22, 1777–1786.

Ratanatharathorn, V., Nash, R.A., Przepiorka, D., Devine, S.M., Klein, J.L., Weisdorf, D., Fay, J.W., Nademanee, A., Antin, J.H., Christiansen, N.P., et al. (1998). Phase III study comparing methotrexate and tacrolimus (prograf, FK506) with methotrexate and cyclosporine for graft-versus-host disease prophylaxis after HLA-identical sibling bone marrow transplantation. Blood 92, 2303–2314.

Scheiermann, C., Kunisaki, Y., and Frenette, P.S. (2013). Circadian control of the immune system. Nat Rev Immunol 13, 190–198.

Schroeder, M.A., and DiPersio, J.F. (2011). Mouse models of graft-versus-host disease: advances and limitations. Dis Model Mech 4, 318–333.

Shlomchik, W.D. (2007). Graft-versus-host disease. Nat Rev Immunol 7, 340–352.

Storb, R., Deeg, H.J., Whitehead, J., Appelbaum, F., Beatty, P., Bensinger, W., Buckner, C.D., Clift, R., Doney, K., Farewell, V., et al. (1986). Methotrexate and cyclosporine compared with cyclosporine alone for prophylaxis of acute graft versus host disease after marrow transplantation for leukemia. N Engl J Med 314, 729–735.

Wang, C., Lutes, L.K., Barnoud, C., and Scheiermann, C. (2022). The circadian immune system. Sci Immunol 7, eabm2465.

Welniak, L.A., Blazar, B.R., Anver, M.R., Wiltrout, R.H., and Murphy, W.J. (2000). Opposing roles of interferon-gamma on CD4+ T cell-mediated graft-versus-host disease: effects of conditioning. Biol Blood Marrow Transplant 6, 604–612.

Zeiser, R., and Blazar, B.R. (2017). Acute Graft-versus-Host Disease - Biologic Process, Prevention, and Therapy. N Engl J Med 377, 2167–2179.

## REFERENCES

1. Sun G, Tang B, Song K, et al. Unrelated cord blood transplantation vs. HLA-matched sibling transplantation for adults with B-cell acute lymphoblastic leukemia in complete remission: superior OS for patients with long-term survival. Stem Cell Res Ther 2022;13(1):500. DOI: 10.1186/s13287-022-03186-3.

2. Harris AC, Young R, Devine S, et al. International, Multicenter Standardization of Acute Graft-versus-Host Disease Clinical Data Collection: A Report from the Mount Sinai Acute GVHD International Consortium. Biology of Blood and Marrow Transplantation : Journal of the American Society For Blood and Marrow Transplantation 2016;22(1) (In eng). DOI: 10.1016/j.bbmt.2015.09.001.

3. Jagasia MH, Greinix HT, Arora M, et al. National Institutes of Health Consensus Development Project on Criteria for Clinical Trials in Chronic Graft-versus-Host Disease: I. The 2014 Diagnosis and Staging Working Group report. Biol Blood Marrow Transplant 2015;21(3):389–401 e1. DOI: 10.1016/j.bbmt.2014.12.001.

4. Kaplan DH, Anderson BE, McNiff JM, Jain D, Shlomchik MJ, Shlomchik WD. Target antigens determine graft-versus-host disease phenotype. J Immunol 2004;173(9):5467–75. (In eng). DOI: 10.4049/jimmunol.173.9.5467.

5. Blazar BR, Taylor PA, McElmurry R, et al. Engraftment of severe combined immune deficient mice receiving allogeneic bone marrow via In utero or postnatal transfer. Blood 1998;92(10):3949–59. (In eng).

